# Automated identification of bolus types in modified barium swallow studies using deep learning: a preliminary study

**DOI:** 10.64898/2026.05.16.26353385

**Authors:** Shitong Mao, Ariana J Sahli, Sheila Nida Buoy, Cayden Hutcheson, Gabriel Andres Gelabert, Carly E.A. Barbon, Mohamed A Naser, Clifton David Fuller, Kristy K. Brock, Katherine A. Hutcheson

## Abstract

**Purpose:** Modified Barium Swallow (MBS) studies utilize videofluoroscopy, a dynamic X-ray technique for evaluating swallowing anatomy and physiology. Each MBS exam typically includes multiple bolus trials, often involving different bolus consistencies. Accurate classification of bolus types is essential, as swallowing dynamics, aspiration risks, and residue levels vary with bolus consistency. In this preliminary study, we propose a deep learning–based approach for automated bolus type classification in MBS, aiming to provide a standardized and efficient framework for automated processing of swallowing assessments.

**Methods:** A total of 206 patients (Mean ± SD age: 60.24 ± 9.02 years; 89.32% men) underwent MBS examinations, comprising 277 individual MBS studies. The dataset included 2,752 bolus-level video segments, categorized by bolus type as follows: 1,711 liquid (IDDSI 0-3, 62.17%), 521 pudding (IDDSI 4, 18.93%), and 520 solid boluses (IDDSI 7, cookie or cracker, 18.89%). To standardize variable video lengths for the data pipeline, each MBS video was temporally segmented into a fixed-length frame sequence, with shorter videos padded using static frames and longer videos randomly cropped to the target length. We employed an Inflated 3D convolutional neural network to develop the deep learning model.

**Results:** Each video segment contained an average of 273.03 ± 195.81 frames. On the independent test set, the deep learning model achieved an overall accuracy of 96.13%, and the macro F1-score was 95.05% in classifying food bolus types within MBS videos.

**Conclusions:** The developed AI-based system demonstrated effective automated classification of food bolus types in MBS videos, representing an important step toward fully automated MBS analysis for swallowing efficiency assessment. The AI model reduces the reliance on manual labels, thereby promising to streamline clinical and research workflows.

## 1. Introduction

The Modified Barium Swallow (MBS) study is a specialized fluoroscopic examination that provides dynamic assessment of swallowing physiology and function(1-3). This critical motor action is a highly complex, coordinated physiological process that plays a fundamental role in human survival by enabling the safe and efficient intake of nutrition and hydration(4-6). When this process is impaired, as in cases of dysphagia (swallowing difficulties), the consequences extend far beyond difficulty with food and liquid ingestion(7, 8). Because the respiratory and digestive systems share a common anatomical pathway at the level of the pharynx, requiring precise neuromuscular coordination to separate swallowing and breathing, dysphagia significantly increases the risk of aspiration, whereby bolus enters the airway(4, 9, 10). This can lead to serious respiratory complications, including recurrent pneumonia, airway obstruction, or chronic pulmonary damage(4, 11).

The MBS study utilizing videofluoroscopy is recognized as the gold standard for assessing swallowing function, oropharyngeal anatomy, and swallowing physiology in real time, as it enables direct visualization of bolus transit as well as the anatomical and physiological processes involved in swallowing(1, 12). It also plays a critical role in diagnosing dysfunction, preventing complications, and guiding therapeutic interventions(3, 13). A standard MBS exam typically involves several bolus trials of different consistencies, since swallowing safety and efficiency may vary substantially depending on bolus type(14, 15). Each bolus trial is generally recorded as a separate video segment, providing a distinct unit of analysis for clinical interpretation or research. The overall interpretation is based on observations of penetration, aspiration, and residue from the oropharynx to the hypopharynx, with explicit consideration of the administered bolus(16). Conducting this procedure is complex: speech-language pathologists (SLPs) must not only instruct patients to perform specific swallowing tasks and evaluate the fluoroscopic images in real-time, but also review, label, and interpret the recorded MBS videos afterward(17-19).

The review process is particularly critical, as it allows clinicians to conduct frame-by-frame analysis and carefully observe subtle changes in bolus transit as well as swallowing safety and efficiency measures(20). However, this process is both labor-intensive and highly dependent on clinical expertise. With the rapid development of deep learning and computational approaches for medical imaging, automated methods have recently been applied to MBS studies to support sophisticated analyses(21). Prior research has demonstrated the promise of deep learning in accurately identifying anatomical landmarks and characterizing bolus transit(19, 22, 23). Nevertheless, to comprehensively evaluate swallowing safety and efficiency, knowledge of the swallowed bolus type or consistency remains indispensable, and this critical factor has not yet been systematically addressed in existing studies(24-26).

For MBS exams, bolus types and volumes vary for each trial during the examination. Best practice aligns the consistencies with the eight-level International Dysphagia Diet Standardisation Initiative (IDDSI) framework (e.g., IDDSI 0 corresponds to thin liquids and IDDSI 7 to solid foods)(27). In routine clinical practice, however, documentation of bolus consistency remains highly variable across institutions. While some reports focus on the consistencies tested and provide overall judgments of swallowing performance, detailed trial-level annotation of bolus consistency is often absent. Moreover, real-time labeling of bolus type during image acquisition is inconsistently implemented. Clinicians may rely on handwritten notes, typed annotations, image markings, or audio recordings to capture this information during the exam, which can be later supplemented in the clinical report. Such supplementary documentation practices implicitly assume that the MBS video itself does not reliably convey the administered bolus type, thereby necessitating external notes to preserve this essential information. The variability and inefficiency of current documentation practices highlight the importance of developing methods that can directly label bolus type information from the recordings. Automatic identification of the administered consistency can reduce the effort needed for the MBS analysis, mitigate documentation errors (that may influence clinical decision making), and substantially improve the documentation process. Moreover, automated identification of food bolus type can be systematically integrated with existing deep learning frameworks for swallowing function assessment, thereby facilitating fully automated MBS analysis and enabling comprehensive and quantitative evaluation.

In this preliminary study, we hypothesize that each bolus trial video inherently contains information about the administered bolus type in MBS, and that this information can be identified through computer vision methods to enable automated bolus type recognition. We proposed to use a deep learning algorithm through a data-driven method that automates the food bolus types. As a preliminary, we focus on the three most used IDDSI levels: liquid (IDDSI 0-3), pudding (IDDSI 4), and solid boluses (IDDSI 7). This investigation provides evidence that the proposed model can effectively categorize different types of swallowing videos, constituting an important step toward the integration of AI-driven approaches in computational deglutition. This work also represents a foundational step toward the investigators’ ongoing effort (P01CA278716) to automate DIGEST (Dynamic Imaging Grade of Swallowing Toxicity) grading(28, 29), which integrates bolus type information at the trial level to generate an overall grade of swallowing safety and efficiency.

## 2. Method

### 2.1 Data collection

In this study, a convenience sample from 206 patients (average age 60.24 ± 9.02 years, including 184 [89.32%] males) at the University of Texas, MD Anderson Cancer Center from 2016 to 2022. The analysis of these MBS videos was approved by the Institutional Review Board (IRB) of the University of Texas MD Anderson Cancer Center (IRB approval number PA19-0261, May 10, 2019). Each patient underwent one or more MBS examinations as part of standard clinical care. All MBS were staffed by licensed-oncology trained SLPs/clinicians within the Section of Speech Pathology and Audiology and administered in the Head and Neck Center at MD Anderson.

The standard MBS protocol typically includes ten bolus trials. In each trial, the patient was instructed by a clinician to swallow a specified volume and consistency of barium material, while the fluoroscopy system acquired and recorded X-ray images in the lateral view, corresponding to the sagittal plane. For each bolus trial, fluoroscopic imaging and video acquisition were typically initiated immediately before bolus administration and terminated immediately after completion of the swallow, rather than being maintained continuously across multiple bolus trials, in order to minimize unnecessary radiation exposure. Thus, each recorded video generally represented a single bolus-trial swallowing event.

The standard MBS protocol includes six thin liquid barium trials, including two 5-mL trials, two 10-mL trials, and two self-administered cup-sip trials using thin liquid barium (IDDSI 0–3); two barium pudding trials (IDDSI 4); and two trials using a cracker coated with barium paste (IDDSI 7). Thickened liquids (IDDSI 1-3) were additionally administered at the discretion of the clinician. All barium sulfate materials used in these trials were Varibar products from Bracco Diagnostics, Inc. (Monroe, NJ). Additional MBS videos may include anterior–posterior view recordings in the coronal plane with thin liquid and/or pudding barium, scout recording obtained as an initial view without bolus administration, and compensatory-strategy trials performed (such as head-turn or chin-tuck) as clinically indicated. No predefined criteria were established for early termination or deviation from the standard MBS protocol; instead, clinicians modified or discontinued the protocol based on clinical judgment when indicated, such as in cases of poor patient tolerance or profound swallowing impairment. The videofluoroscopy in MBS was performed using Siemens Luminos dRF Max or Artis Zee digital fluoroscopy system (Siemens Healthineers, Forchheim, Germany). The swallowing videos were acquired at 30 frames per second (FPS). The video stream was recorded using TIMS MVP (TIMS Medical, Chelmsford, MA, USA) and exported in DICOM format with synchronized audio when available. All individual video frames were acquired as grayscale images. All patient information containing direct identifiers and names, was de-identified before model development.

In this study, 277 MBS studies from 206 patients were included. In this study, 277 MBS studies from 206 patients were included. Clinicians labeled the bolus trial in real-time in the fluoroscopy suite using the TIMS MVP labeling feature. For this study, the videos were independently reviewed by two additional raters, who annotated the bolus type for each swallowing trial according to its consistency. Videos were excluded if they met any of the following criteria: 1. Anterior–posterior view recordings of bolus trials; 2. Scout recordings, defined as videos without actual bolus administration or swallowing; 3. Videos for which the two independent raters assigned different bolus-type labels and, after adjudication by a third rater, the bolus type remained problematic or could not be reliably determined (<0.1%); 4. Videos for which at least one of the two raters labeled the bolus type as “uncertain” and, after adjudication by a third rater, the bolus type remained problematic or could not be reliably determined (<0.25%); 5. Other tested food consistencies, such as sauce/minced fruit (IDDSI 5) or soft diced fruit (IDDSI 6). Of note, videos excluded under criteria 3 and 4 were likely to include these food consistencies. Swallowing videos involving compensatory strategies were not excluded from the dataset. During bolus-type annotation, raters classified each trial solely according to bolus consistency, independent of swallowing physiology or impairment-related findings, including penetration, aspiration, or pharyngeal residue severity. A final dataset of 2,752 swallowing videos was retained for analysis. Based on consistency annotation, 1706 videos were classified as liquid bolus trials (IDDSI 0–3; 62%), 523 as pudding bolus trials (IDDSI 4; 19%), and 523 as solid bolus trials (IDDSI 7; 19%). This distribution was approximately consistent with the bolus-type distribution expected under the standard clinical MBS protocol, in which liquid, pudding, and solid bolus trials are typically administered at a ratio of 3:1:1.

### 2.2 Data pipeline

For the subsequent deep learning–based bolus-type classification task, all videos were required to be represented as fixed-length frame sequences before model input. Given that the original MBS swallowing videos varied substantially in duration across bolus trials, ranging from 50 to 1,740 frames (Results), a temporal standardization procedure was applied using a predefined number of frames per video (NFPV) as the target sequence length, as shown in Fig. 1. For videos shorter than NFPV, temporal padding was applied by replicating the first frame before the original sequence and the last frame after the original sequence until the target length was reached. The number of frames padded at the beginning and the end was randomly determined. For videos with a length equal to or greater than NFPV, we applied two temporal sampling strategies: fixed-window and uniform-sampling. In the fixed-window strategy, a continuous segment containing NFPV frames was randomly selected from the original video. In the uniform-sampling strategy, NFPV frames were sampled at approximately equal intervals across the entire video duration. Those two strategies ensured that all videos were formatted as fixed-length inputs while preserving the temporal structure of each bolus-trial swallowing event.

**Fig. 1.**
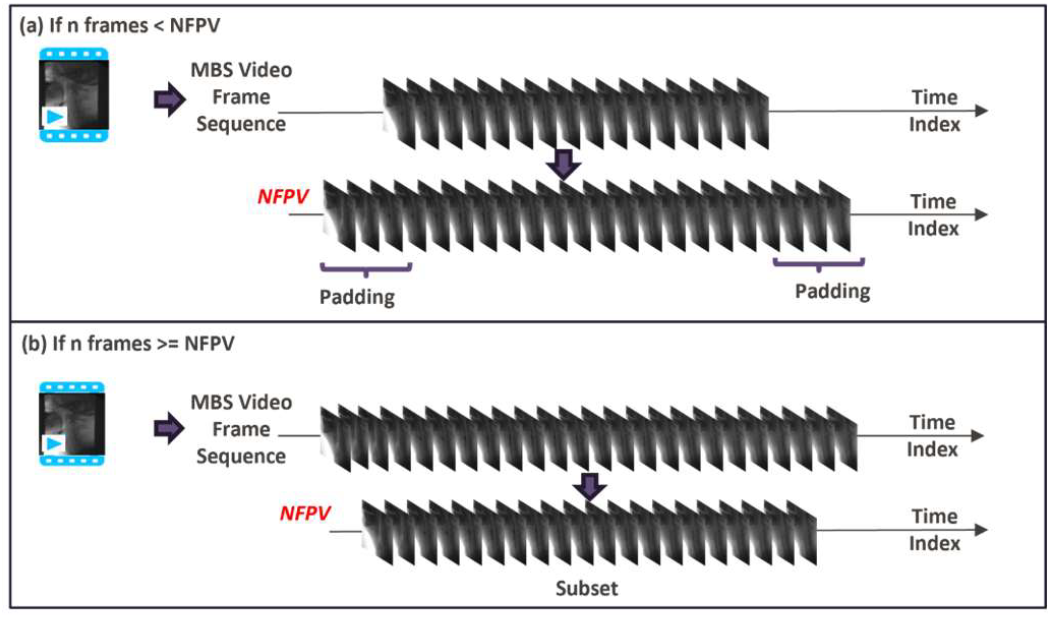
Temporal standardization of MBS videos using frame padding for shorter videos and subset selection for longer videos.

### 2.3 Training and validation

In this study, we followed a cross-subject prediction approach for designating training, validation, and testing sets as outlined in(30), Specifically, all MBS videos collected from the same patient were assigned to the same dataset partition and were not split across the different sets. The data were split into training, validation, and testing sets at approximately 70%, 15%, and 15%, respectively. Details on data distribution are outlined in Table I. The gender distribution was attributable to the clinical cohort characteristics, as the majority patients in this study were diagnosed with head and neck cancer, which has been well-investigated to exhibit a higher incidence in male individuals(31, 32).

**Table I.**
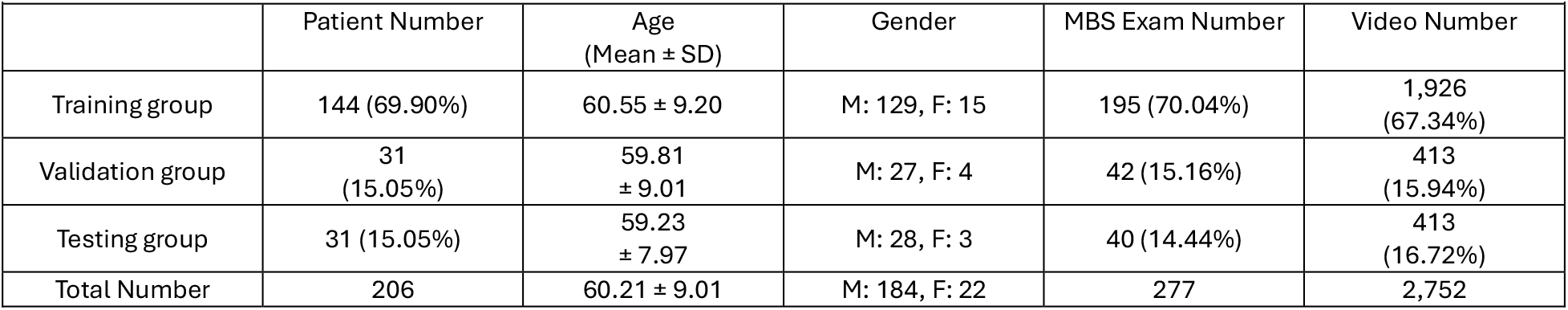
The demographic distribution for the training, validation and testing sets.

Model optimization was performed using the training set, and model performance was monitored on the validation set after each epoch. Early stopping was applied by terminating training if validation accuracy did not improve for 50 consecutive epochs. The model checkpoint with the highest validation accuracy was selected as the final model and subsequently evaluated on the independent testing set. The random temporal padding or segment selection procedure was applied during training and validation. For final evaluation on the independent testing set, the same temporal standardization procedure was used, with each testing video evaluated five times and the final predicted bolus type determined by majority voting across the five predictions.

### 2.4 Other setups

We used an Inflated 3D Convolutional Network (I3D) model (with a ResNet-50 backbone) for video-based bolus-type classification(33). Considering computational resource constraints, we implemented a single-stream architecture using only the video input stream, rather than the original two-stream design that additionally incorporates optical flow(34). Because all MBS video frames were originally grayscale images (single-channel images) and the I3D model requires a three-channel image input, each MBS grayscale frame was converted into a three-channel format by duplicating the same grayscale image across all three channels. This preprocessing step allowed the grayscale MBS videos to be used as model input without altering the original image content. The I3D model was initialized with pretrained weights from PyTorchVideo, which were trained on the Kinetics-400 human action recognition dataset(35) and then fine-tuned on the MBS data.

To improve model robustness, data augmentation was applied to the training set during model training. For each sampled training video, random rotation was performed within a range of −30° to 30°. Random horizontal flipping was applied with a probability of 50%, whereas vertical flipping was not used to preserve the upright patient orientation typical in MBS exams. Random cropping was also applied, with the cropped region covering 80% to 100% of the original frame area and the width-to-height aspect ratio randomly selected between 0.6 and 1.667. All frames were resized to 256 × 256 pixels before model input. We employed accuracy (ACC) and macro-averaged F1 (MF1), as the primary metrics to assess model performance on the testing set. Training employed a Stochastic Gradient Descent (SGD) optimizer (learning rate=0.0001, momentum=0.9). To address class imbalance across bolus-type categories (0.62:0.19:0.19), model training was performed using a weighted cross-entropy loss function. During model training, the batch size was adjusted according to the NFPV setting: a batch size of 16 was used when NFPV was set to 32 or 64, whereas a batch size of 8 was used when NFPV was set to 128. This study was conducted on a Kubernetes GPU cluster, where each node had 128 physical CPUs, 1 TB of memory, and 8 Nvidia A100 GPUs.

## 3. Results

### 3.1 Dataset Characteristics

The image characteristics of the collected MBS videos were summarized to characterize the variability of the input data, as shown in Fig. 2. Video resolution varied across the MBS dataset, with the two most common resolutions being 1024 × 952 pixels and 1280 × 998 pixels, accounting for 27.8% and 26.1% of videos, respectively. Other frequently observed resolutions included 1280 × 1024 pixels (9.8%), 956 × 1024 pixels (8.4%), and 1024 × 1024 pixels (8.7%), whereas the remaining resolutions each accounted for a smaller proportion of the dataset.

**Fig. 2.**
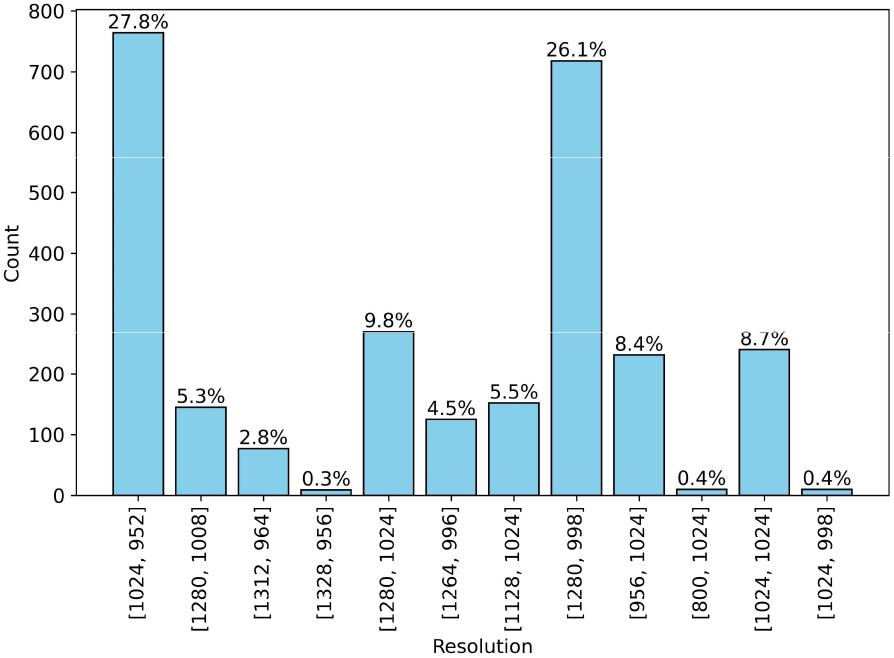
Distribution of spatial resolutions across MBS swallowing videos.

The distribution of video length is shown in Fig. 3. Video length varied substantially, ranging from short recordings (50 frames) to markedly longer videos (1740 frames). The mean value of the frames was 273.03 and the standard deviation was 195.81. The red curve in Fig. 3 represents the kernel density estimate of the distribution, with its peak at 161.65 frames, indicating the most common video length range.

**Fig. 3.**
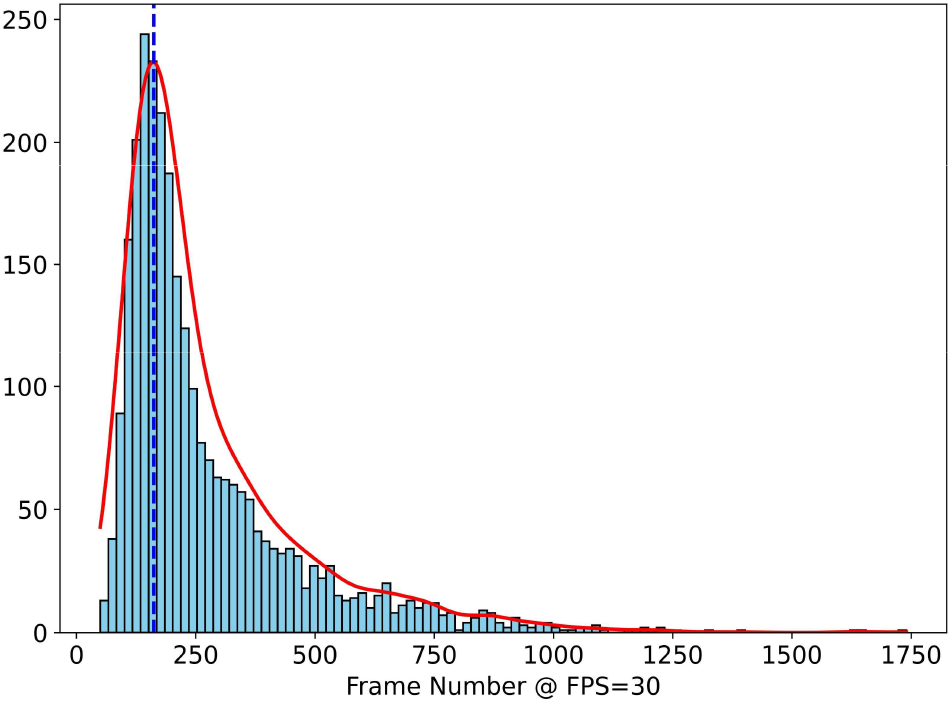
Distribution of video length measured by frame count at 30 FPS.

### 3.2 Classification Performance

To evaluate the efect of input preprocessing on model performance, we examined diferent strategies for defining the fixed-length video input, fixed-window and uniform-sampling, as described in 2.2. We further compared model performance across diferent fixed-length input settings, with the number of frames per video (NFPV) set to 32, 64, and 128. Also, we compared two approaches for pixel-value normalization. The original MBS video frames had grayscale intensity values ranging from 0 to 255. In the first normalization approach, frame intensities were rescaled to the range of 0 to 1 before model input. In the second approach, after rescaling to 0 to 1, the frames were further normalized using the standard preprocessing parameters applied to I3D models for conventional color video inputs, in which each pixel value was centered by subtracting 0.45 and scaled by dividing by 0.225. The ACC and MF1 are listed below:

In Table II, under 0–1 intensity rescaling, fixed-window sampling increased from 91.04% accuracy and 88.95% MF1 at NFPV = 32 to 95.41% accuracy and 94.16% MF1 at NFPV = 128. A similar trend was observed with color intensity normalization. For uniform sampling, increasing NFPV from 32 to 64 slightly improved performance, whereas further increasing NFPV to 128 reduced both accuracy and MF1. Uniform sampling with 0–1 intensity rescaling achieved the best performance, with the highest accuracy of 96.13% and the highest MF1 score of 95.05% when NFPV was set to 64. In contrast, the fixed-window strategy showed lower performance at smaller NFPV values, although its performance improved as NFPV increased.

**Table II.** Model performance across temporal sampling strategies and intensity normalization approaches settings.

|  |  |  |  |  |
| --- | --- | --- | --- | --- |
|  | NFPV | 32 | 64 | 128 |
| Fixed-window +<br>0-1 intensity rescaling | ACC | 91.04% | 93.22% | 95.41% |
|  | MF1 | 88.95% | 90.36% | 94.16% |
| Fixed-window +<br>Color intensity<br>normalization | ACC | 90.80% | 93.22% | 94.19% |
|  | MF1 | 88.26% | 91.10% | 92.12% |
| Uniform sampling +<br>0-1 intensity rescaling | ACC | 95.88% | **96.13%** | 93.46% |
|  | MF1 | 94.46% | **95.05%** | 90.73% |

## 4. Discussion

This preliminary study demonstrated the feasibility of using advanced deep learning methods to automatically identify bolus type at the bolus-trial level in MBS videos. Bolus consistency represents a fundamental element that must be determined during MBS analysis and provides essential clinical context for interpreting swallowing performance. The clinical relevance of penetration, aspiration, or pharyngeal residue may differ depending on whether the swallowed bolus is thin liquid, pudding, or solid. Therefore, automated bolus-type identification promises to help subsequent clinical workflows evaluate swallowing findings within the appropriate bolus-specific context. This approach can be computationally integrated with well-developed deep learning–based algorithms for MBS analysis, including imaging-based anatomical landmark detection and bolus segmentation. Together, these tools could provide complementary outputs within a more comprehensive computational framework for MBS analysis, supporting clinicians in the observation of potential swallowing impairment and subsequent clinical decision-making. Importantly, determining bolus type is an essential component of assessing swallowing efficiency using the investigators’ DIGEST framework. Automating bolus-type identification through artificial intelligence may therefore represent an important step toward automated DIGEST grading.

Additionally, this study also demonstrated the feasibility of using intrinsic video-derived information to identify bolus type. Conventional documentation methods in Modified Barium Swallow (MBS) studies implicitly assume that the videofluoroscopic recordings themselves do not contain information about the administered bolus type or that such information is not readily distinguishable by visual inspection alone. As a result, clinicians must rely on supplementary approaches such as audio labeling, during the exam, manual tagging in swallow recording systems, or handwritten notes to supplement this information for clinical record or retrospective review. However, these strategies vary widely across institutions and are rarely standardized, leading to inconsistencies in how bolus types are recorded and reported. This variability introduces a source of human error or ambiguity in clinical interpretation and threatens reproducibility in research settings. Automated identification of bolus types directly from MBS recordings offers a means to overcome this limitation by embedding the bolus information into the video data itself, thereby reducing reliance on manual documentation and promoting consistency across clinical and research environments.

A major methodological challenge in this study was addressing the substantial variability and excessive length of the original MBS videos, whereas the deep learning model required fixed-length video inputs. This issue is less prominent in many prior deep learning applications for video classification, such as human action recognition, where videos are more standardized or already centered around a target action. The Kinetics-400 dataset consists of video clips of approximately 10 seconds, and the original I3D study using Kinetics, models were trained using 64-frame snippets(35). The length variability observed in MBS videos was substantially greater than that typically encountered in Kinetics-400, making direct use of the full video sequence impractical for model input. Therefore, a temporal standardization strategy was required to reduce each video to a fixed number of input frames. In our experiments, temporal uniform sampling consistently outperformed fixed-window sampling based on continuous frame segments. This finding suggests that, for MBS videos with substantial variation in duration, selecting only a continuous fixed-length segment may skip clinically relevant information occurring outside the sampled window. In contrast, uniform sampling across the full video duration may better preserve global temporal information from the entire swallowing event. Under the uniform sampling strategy, NFPV = 64 achieved better performance (ACC=96.13%, MF1= 95.05%) than NFPV = 128 (ACC=93.46%, MF1= 90.73%). One possible explanation is that increasing the number of sampled frames to 128 introduced more redundant or less informative frames, thereby increasing the learning difficulty and adding temporal noise to the model input. In addition, the smaller batch size required for NFPV = 128 (batch size = 8) may have reduced training stability. By contrast, NFPV = 64 may better match the effective temporal scale of the swallowing event, providing sufficient temporal coverage while maintaining a higher concentration of task-relevant information.

Another potential challenge was the high degree of background similarity across MBS videos. Because patients were positioned in a similar seated posture and imaged in a consistent field of view, the videos shared highly similar grayscale backgrounds, while the bolus types were different. This characteristic differs substantially from many conventional video classification datasets, in which background, color, viewpoint, and scene content are typically more variable. Despite this challenge, the experimental results indicate that the selected I3D model without optical flow was able to distinguish bolus-related dynamic features from videos with highly similar backgrounds. Variability in image resolution could potentially introduce spatial deformation after random cropping and resizing, which may further modify the appearance of anatomical structures and bolus patterns. Nevertheless, the experimental results suggest that this was not a primary source of model error. One possible explanation is that the range of resolution variation in the dataset was relatively limited, allowing the resizing procedure to standardize the input frames without substantially distorting task-relevant visual information. MBS videos consist of grayscale medical images, and it was initially unclear whether an I3D model pretrained on conventional color videos would be sensitive to the choice of intensity normalization. Therefore, two normalization approaches were evaluated in this study. The experimental results showed that the difference between these normalization strategies had limited influence on final classification performance.

This study has several limitations. First, all patients included in this study were head and neck cancer patients, therefore, the performance of the model in dysphagia populations with other clinical situations, such as neurodegenerative disease, remains unclear. Second, the bolus type categories used in this study were defined based on MD Anderson Cancer Center’s institutional standard MBS protocol and the bolus consistencies most relevant to future automated DIGEST assessment. As a result, other food consistencies or less commonly administered bolus types were not included in the current classification framework. For this reason, future work is needed to extend this model to discriminate against other consistencies. From a technical perspective, video-based deep learning classification is computationally demanding. Although the available computational resources for this study were substantially strong, they did not allow the use of larger batch sizes, particularly for longer input sequences. This constraint may have introduced additional variability during model optimization. In addition, we implemented a single-stream I3D architecture without optical flow because of computational resource constraints. Although the model achieved accuracy greater than 96%, it remains unknown whether incorporating an additional optical-flow stream would further improve classification performance.

## Conclusion

Bolus consistency provides essential context for evaluating swallowing performance and is a required component for DIGEST-based assessment of swallowing efficiency. Automated bolus-type recognition represents an important step toward more comprehensive AI-assisted MBS analysis. In this study, the proposed deep learning-based system demonstrated the feasibility of automatically classifying bolus type from MBS swallowing videos. This approach may support future integration with automated landmark detection, bolus segmentation, and swallowing safety and efficiency assessment, ultimately enabling a more standardized and scalable computational workflow for MBS interpretation.

## Data Availability

All data produced in the present study are available upon reasonable request to the authors

## Acknowledgements

We would like to express our sincere appreciation to Emory Twitty, Viridiana Andrade, and Parvin Ebadi, for their expert and dedicated manual annotation of the videos.

